# Protocol of a Study to Benchmark Occupational Health and Safety in Japan: W2S-Ohpm Study

**DOI:** 10.1101/2023.03.11.23287146

**Authors:** Tomohisa Nagata, Kiminori Odagami, Masako Nagata, Nuri Purwito Adi, Koji Mori

## Abstract

We aim to conduct a prospective cohort study to benchmark occupational health and safety in Japan. Here, we describe the detailed protocol for the baseline survey based on the Checklist for Reporting Results of Internet E-Surveys. We conducted the baseline survey for the prospective cohort study in 2022. Our target population was workers in Japan aged 20 years or older, who we sampled to be representative of the Japanese workforce, stratified by sex, age, and region. Among 59,272 registered monitors who answered the initial screening questions, 29,997 completed the survey. After excluding 2,304 invalid responses, we used 27,693 valid participants in our final analysis. The number and mean age of men were 15,201 (55%) and 46 years; those of women were 12,492 (45%) and 45 years. With respect to sex, age, and regional composition, our sampling was representative of Japan’s working population. Our sampling for employment status and industry yielded almost the same proportions as a government-led representative sampling of workers in Japan.

## Introduction

Occupational health and safety should be managed continuously by improvements based on the goals and plans of management systems at both the company and national levels. The International Labour Organization (ILO) Convention No. 187 was established in 2006 (1). Its Article 5 states as follows: “Each member shall formulate, implement, monitor, evaluate and periodically review a national programme on occupational safety and health in consultation with the most representative organizations of employers and workers.” As the first country to adopt that convention as a standard, Japan has operated the Occupational Safety and Health Program since 1958 (2). That program is in the form of a 5-year plan, and it is reviewed in accordance with ongoing developments. Such reviews demand highly accurate statistical details of occupational health and safety activities as well as occupational accidents.

Japan’s government regularly obtains information about occupational health and safety. Investigations into occupational safety and health in that country have been conducted as general statistical surveys under the Statistics Law (3). To determine the current situation and ensure nationwide representativeness, we adopted a stratified sampling method. Those government surveys have the advantage of large sample size and random countrywide sampling; however, there are drawbacks, such as the enormous survey costs and the fact that many are cross-sectional—not longitudinal—studies. A cross-sectional study makes it difficult to determine causality because time-series relationships cannot be included in the analysis.

Online surveys are becoming more widely used in research. Many concerns have been raised about the quality of Web-based surveys (4). However, Internet surveys have the advantage of a large sample size at relatively low cost compared with paper-based surveys. Studies using online panels have been widely applied because they are inexpensive and allow for data acquisition over a short period of time (5). Surveys using online panels can be applied to provide follow-up information, thereby permitting longitudinal studies, such as prospective cohorts. In online panel surveys, sampling methods are important, and respondent bias has to be assessed.

We conducted a prospective cohort study on occupational health and safety in Japan. This paper describes the detailed protocol for that study. We report on the research protocol using the Checklist for Reporting Results of Internet E-Surveys (CHERRIES), which is a means for improving the quality of Web surveys (6).

## Materials and Analysis

This is a prospective cohort study conducted online. We undertook the baseline survey for the study from February 28 to March 3, 2022; the follow-up surveys will be conducted annually. The target population for the survey was workers in Japan aged 20 years or older. The study was approved by the Ethics Committee of the University of Occupational and Environmental Health, Japan (R3-076).

### Sampling Plan

To ensure that the target population was representative of workers in Japan, the participants were sampled so that the proportion of workers stratified by sex, age, and region was the same as the actual Japanese workforce (7). We categorized as follows: sex into two groups (men, women); age into eight groups (20–29, 30–39, 40–44, 45–49, 50–54, 55–59, 60–64, and _≥_65 years); and region into 10 groups listed from north to south (Hokkaido, Tohoku, South Kanto, North Kanto and Koshinetsu, Hokuriku, Tokai, Kinki, Chugoku, Shikoku, and Kyushu and Okinawa). We set the target sample size at around 30,000 participants, based on a previous study (8). We assigned the number of workers in each of the 160 collection units stratified by sex, age and region, and the sample size for each collection unit as indicated in Table 1.

**Table 1.**
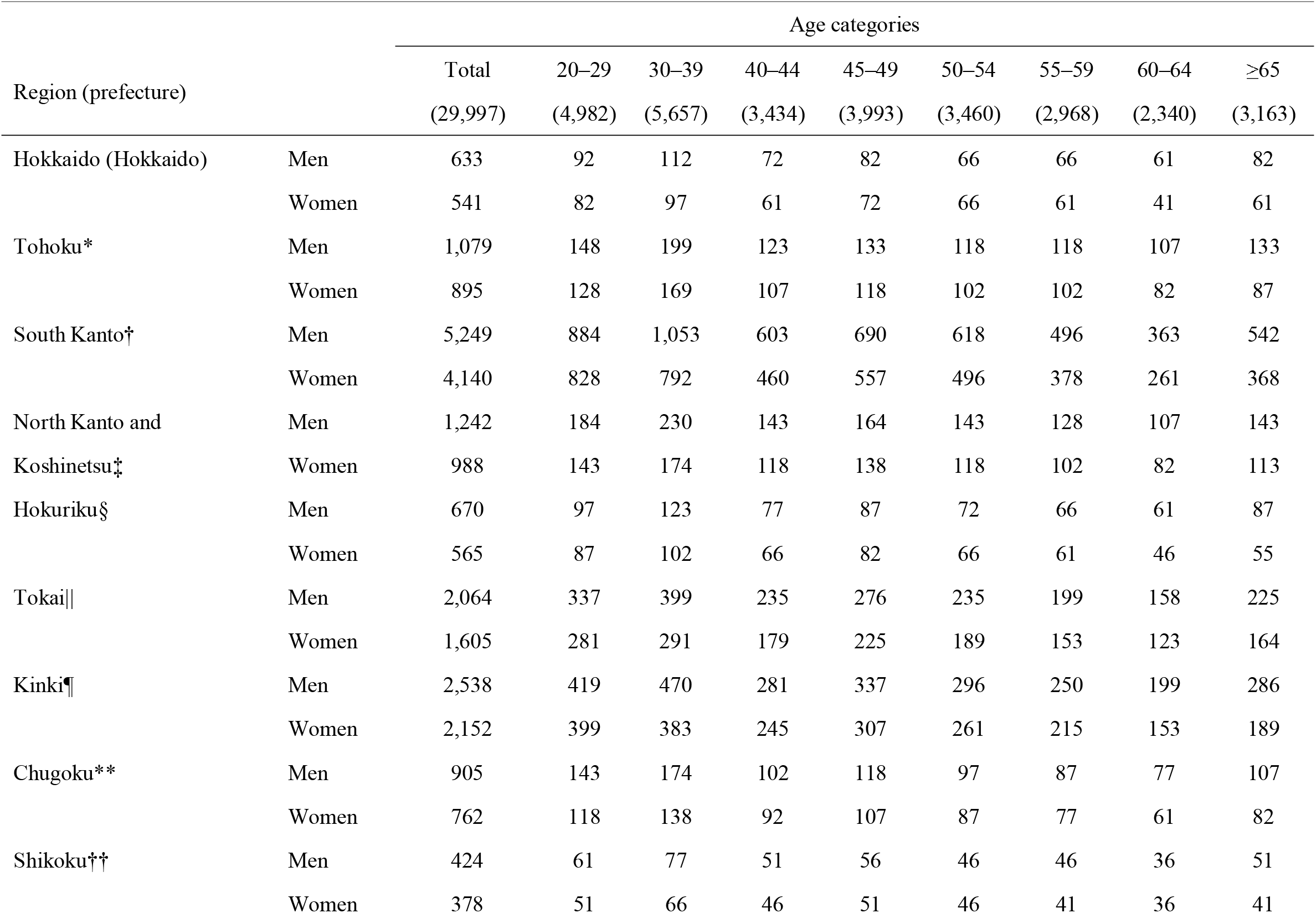
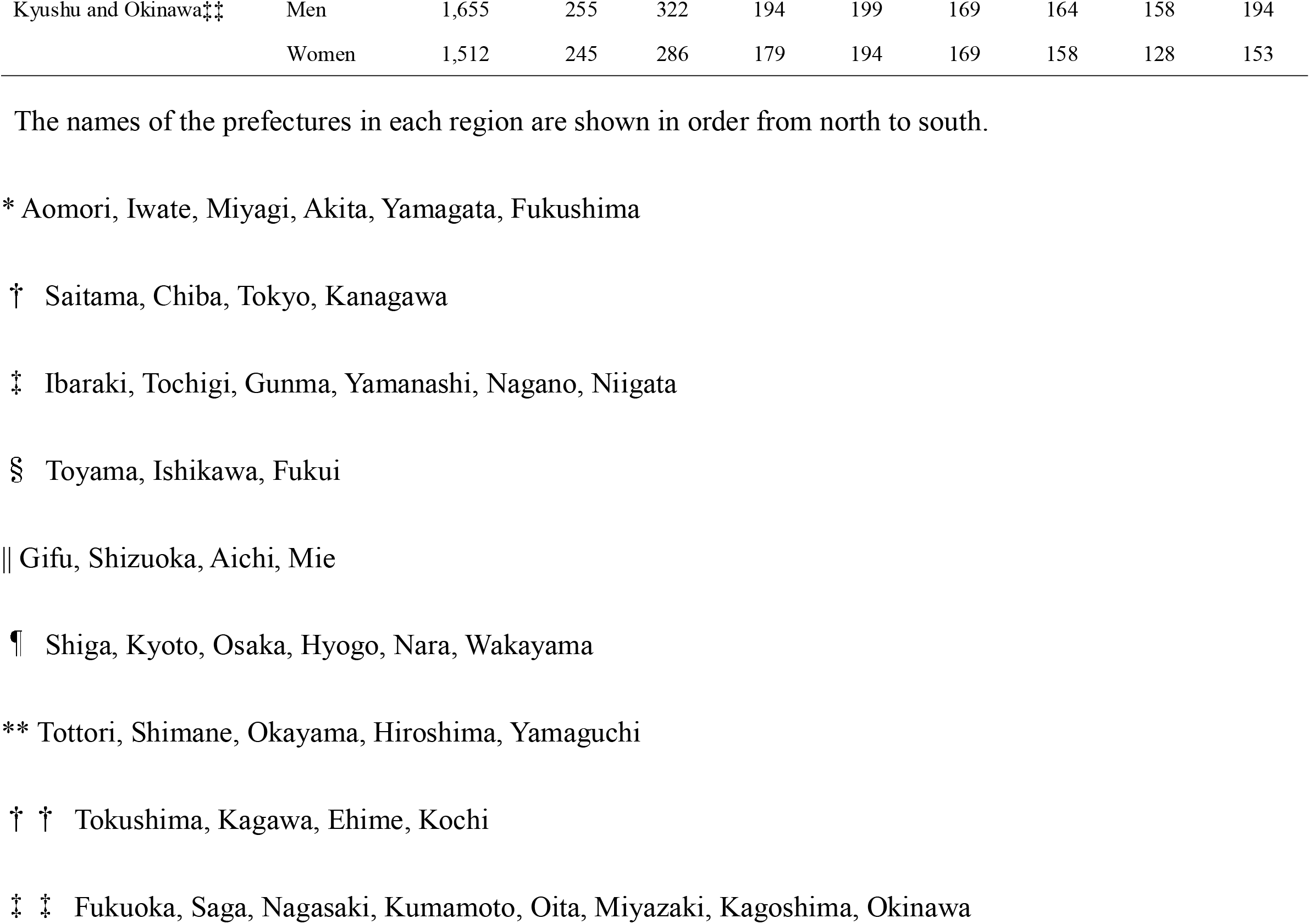
Participants stratified by sex, age, and region in the sampling plan

### Recruitment Procedure

The survey was commissioned by Rakuten Insight, Inc. (Tokyo, Japan), which has 2.2 million registered monitors. Only individuals registered with Rakuten Insight could respond to the Internet survey. Owing to the company’s information confidentiality, we were unaware of the number of initial invitations to participate; however, 59,272 registered monitors responded to the initial screening questions and participated in the survey. Those 29,997 matched the survey’s criteria (worker status, sex, age, and region). As an incentive, survey respondents were able to earn redeemable points from the company; however, again owing to the company’s information confidentiality, we did not know the number of points.

### Data Retrieval

Before submitting the questionnaire, we undertook consistency or completeness checks. For inconsistency in a question, respondents were asked to view on-screen indications that their answers were inconsistent; specifically, that comprised individuals who provided meaningless characters and symbols in the free-answer column (e.g., “AAA” and “$&¥”). This approach has been shown to be effective in detecting fraudulent responses at an early point in surveys (9). We also excluded participants who responded in an excessively short period of time: we regarded such responses as invalid. As post hoc means to eliminate fraudulent responses, we set the exclusion criteria as follows: extremely high body weight (>300 kg); excessive height (>250 cm); and clearly incorrect answers (respondents who indicated that they were engaged in work for 0 days and 0 hours; those who stated they worked over 150 hours a week; those who did over 100 hours overtime a week; and those who claimed they lived with 18 or more family members).

### Measurements

The survey items included basic sociodemographic characteristics, such as sex, gender, nationality, marriage status, income, employment status, and industry category. “Sex” signified biological sex and was determined as the sex on the family register at birth and birth certificate, whereas “gender” was a self-identified term. To identify “gender,” we asked the question, “Regardless of your sex at birth, please indicate the gender that most closely matches your current perceived image”; respondents chose from three options (male, female, and other). Marital status was classified into three categories: married; unmarried; and divorced or bereaved. We determined household income (total income of all cohabiting family members) over 1 year (2021), with options given in units of 1 million Japanese Yen. We classified household income into the following six categories: <4 million; 4 million–6 million; 6 million–8 million; 8 million–10 million; 10 million–12 million; and >12 million Japanese Yen. Respondents were asked to select their employment status from the following eight options: self-employed; company executive; full-time employee; part-time work; dispatched employee; contract employee; freelance; and other. As noted earlier, we excluded non-working individuals from this study at the screening stage.

For comparison with various surveys conducted by the Japanese government, we classified industries into the following 20 categories based on the Japan Standard Industrial Classification (10): agriculture and forestry; fisheries; mining and quarrying of stone and gravel; construction; manufacturing; electricity, gas, heat supply and water; information and communications; transport and postal services; wholesale and retail trade; finance and insurance; real estate and goods rental and leasing; scientific research, professional and technical services; accommodations, eating and drinking services; living-related and personal services and amusement services; education and learning support; medical, health care and welfare; compound services; services; public sector; and unlabeled. This classification is roughly—though not perfectly—consistent with the International Standard Industrial Classification (11); thus, we believed it would allow international comparisons to be made.

We examined negative health conditions through psychological distress using Kessler 6 (K6) as well as diseases and their treatment status. K6 was developed to screen for psychological distress (12, 13), and the Japanese version has been validated (14, 15). Based on the previous studies, we applied a K6 score of 5 or higher as the cutoff for mild psychological distress and one of 13 or higher as the cutoff for severe psychological distress. Positive health conditions were examined by work engagement using a nine-item Japanese version of the Utrecht Work Engagement Scale (UWES-9) (16, 17). The total score was obtained by averaging the individual item scores (possible range, 0–6). The internal reliability and validity of the Japanese UWES-9 are acceptable (17).

### Statistical Analysis

We have indicated the number of participants for analysis and those we judged to have given invalid responses by sex and age category. We compared the differences between those two groups using *t* tests for continuous variables (such as age, K6, and work engagement score) and chi-square tests for categorical variables (such as sex and nationality). We determined age, gender, nationality, marital status, household annual income, employment status, industry, K6, and work engagement score by sex for the 27,693 participants who excluded invalid responses.

## Results

We were able to obtain the desired number of participants for sex, age and regional stratification. Detailed information based on the CHERRIES appears in Table 2.

**Table 2.**
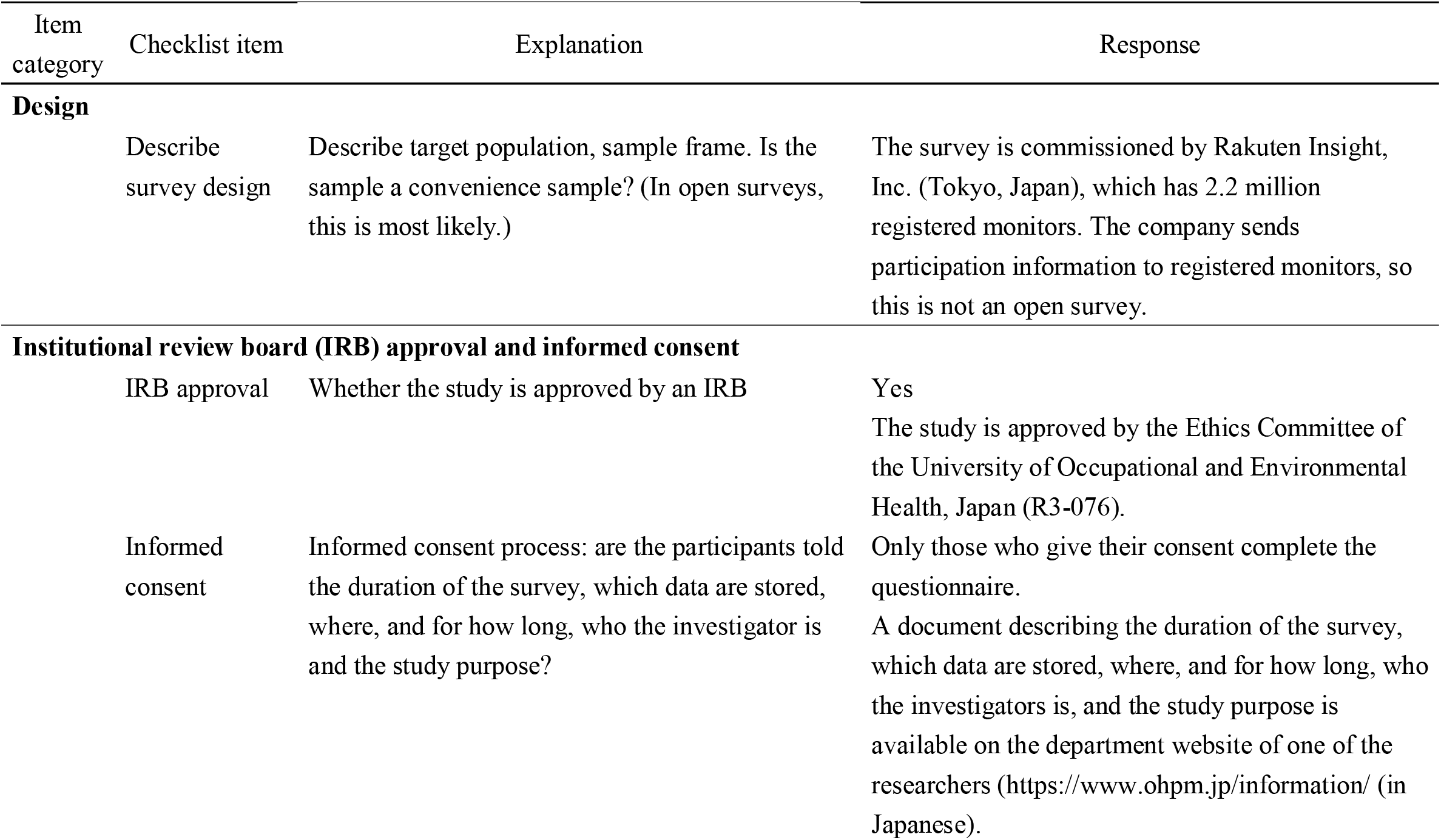
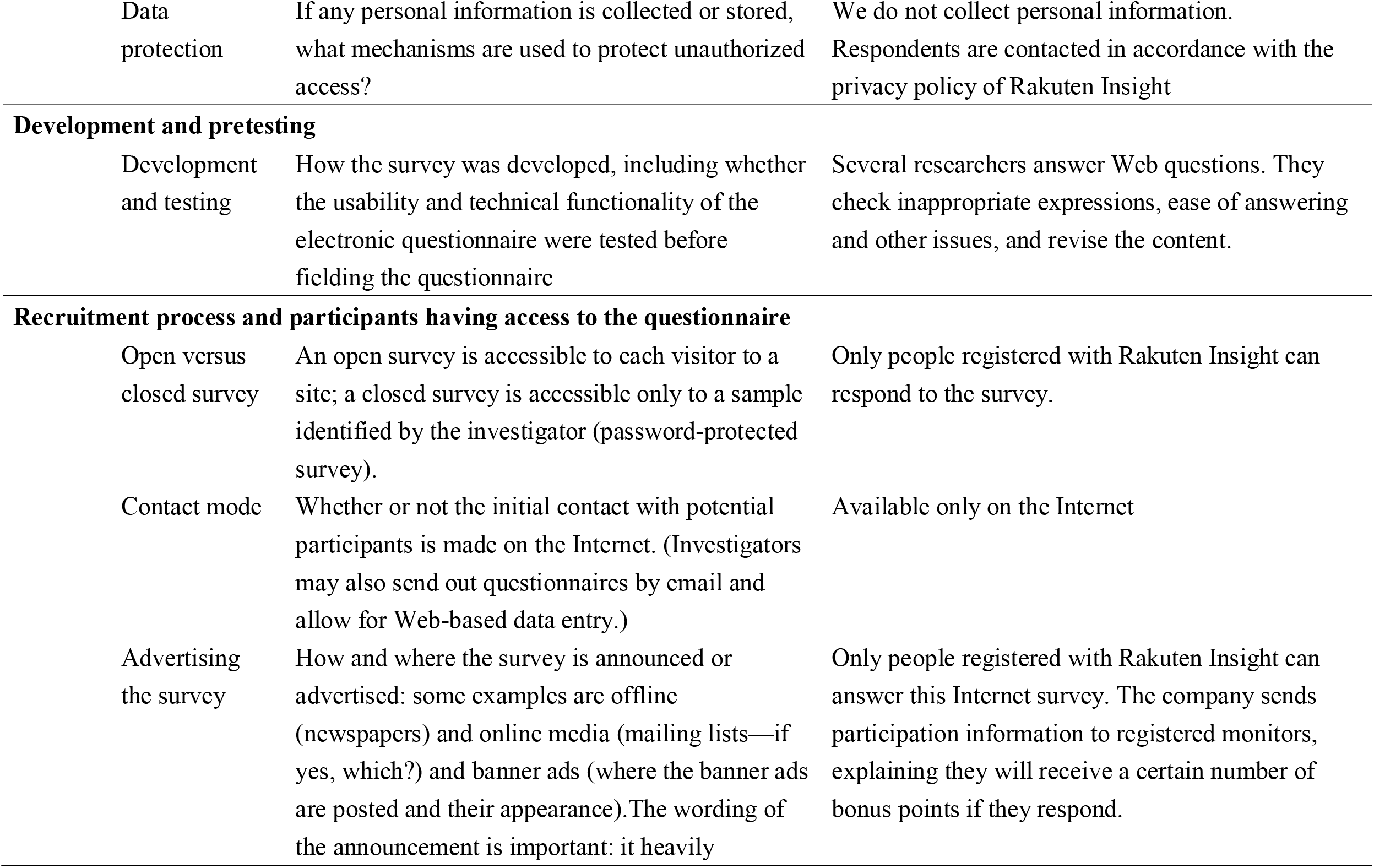
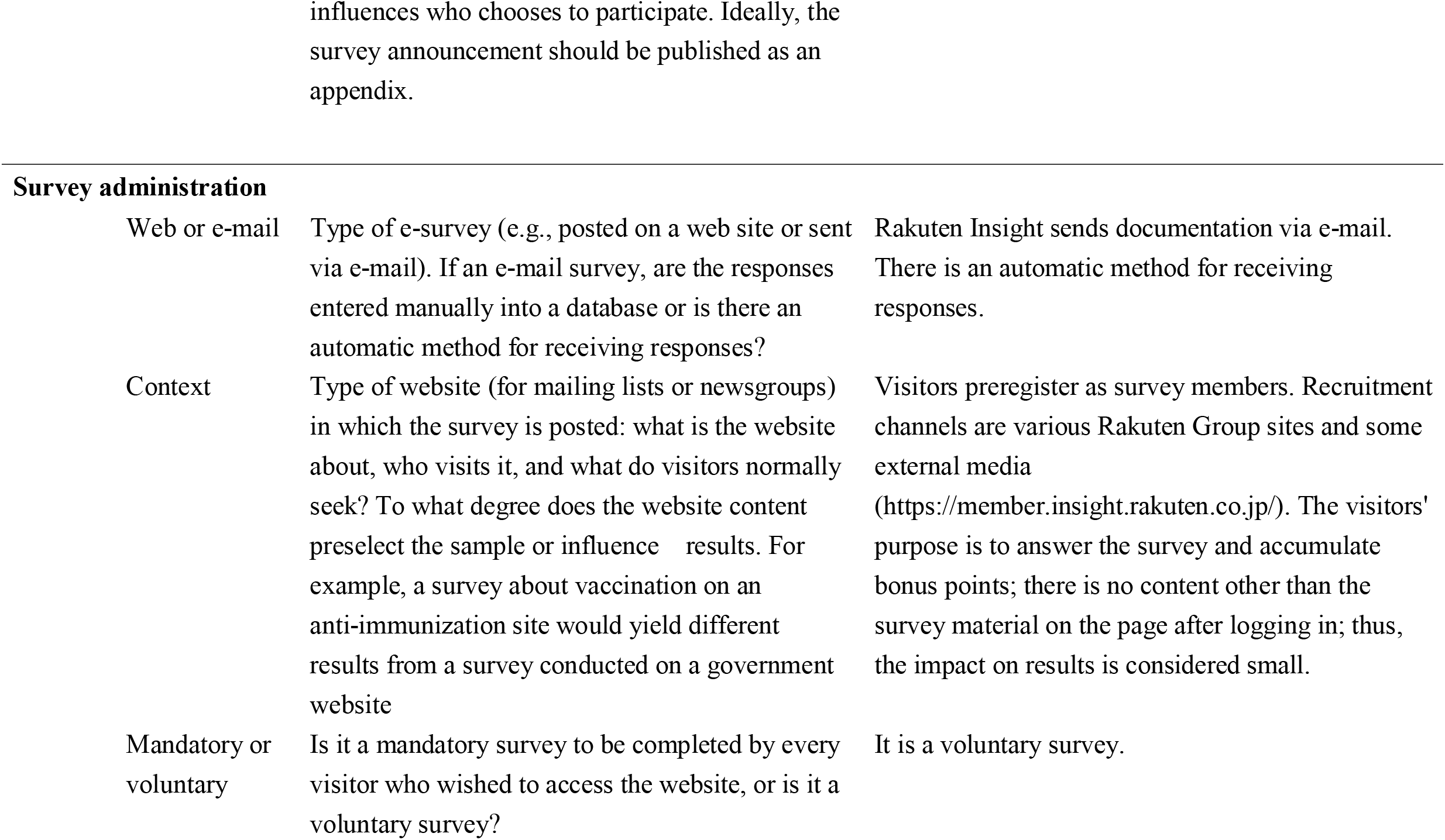
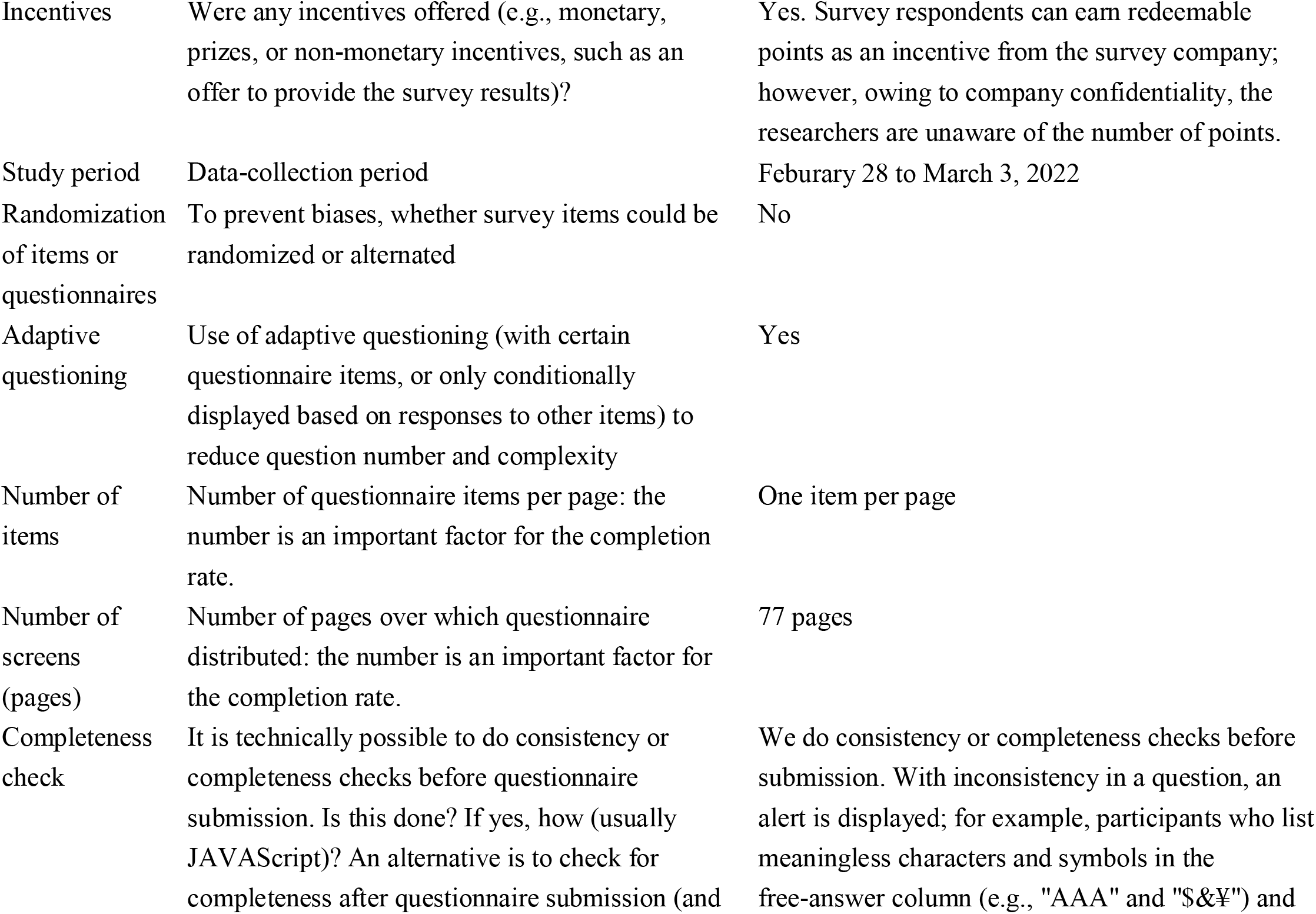
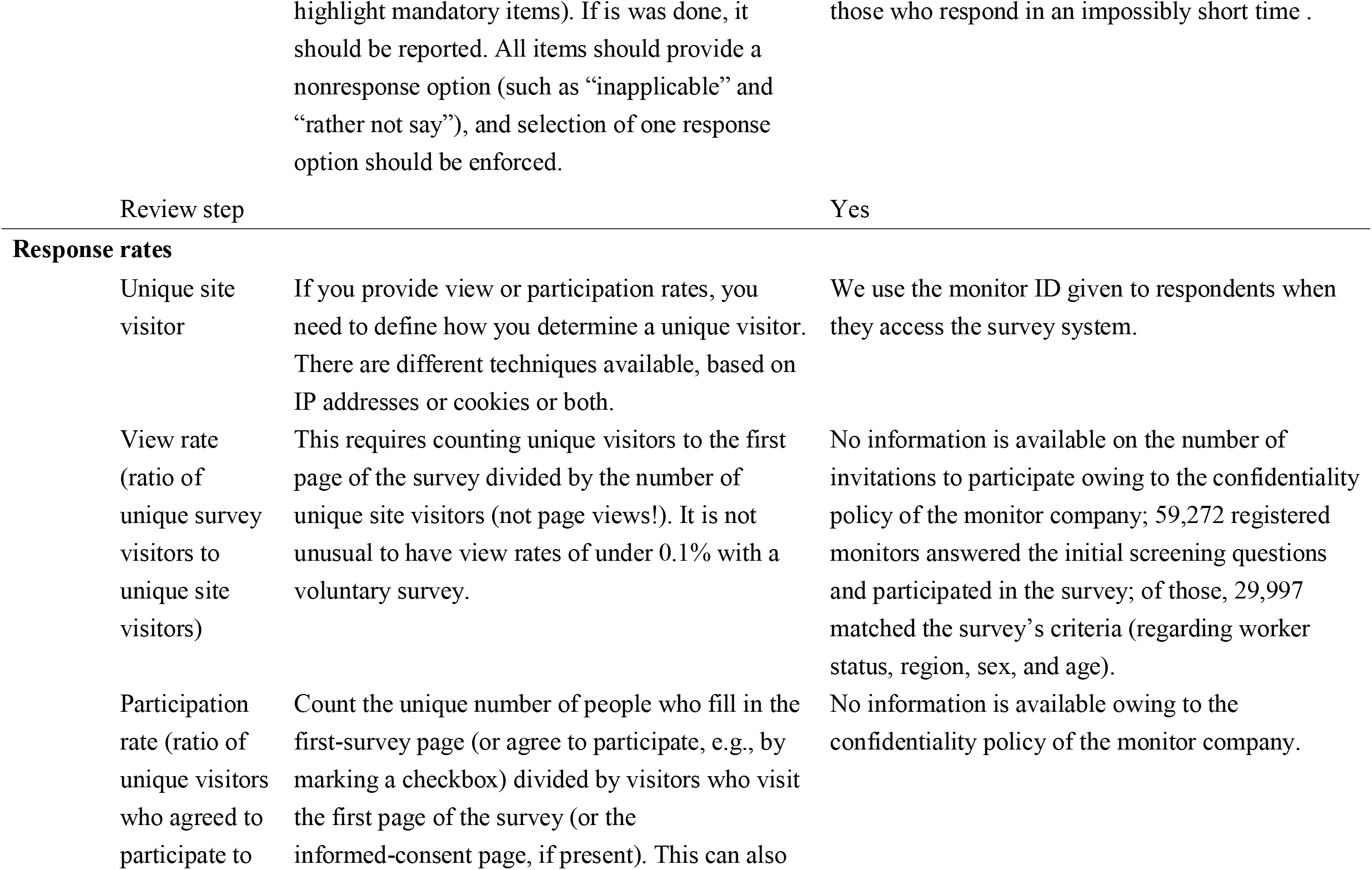
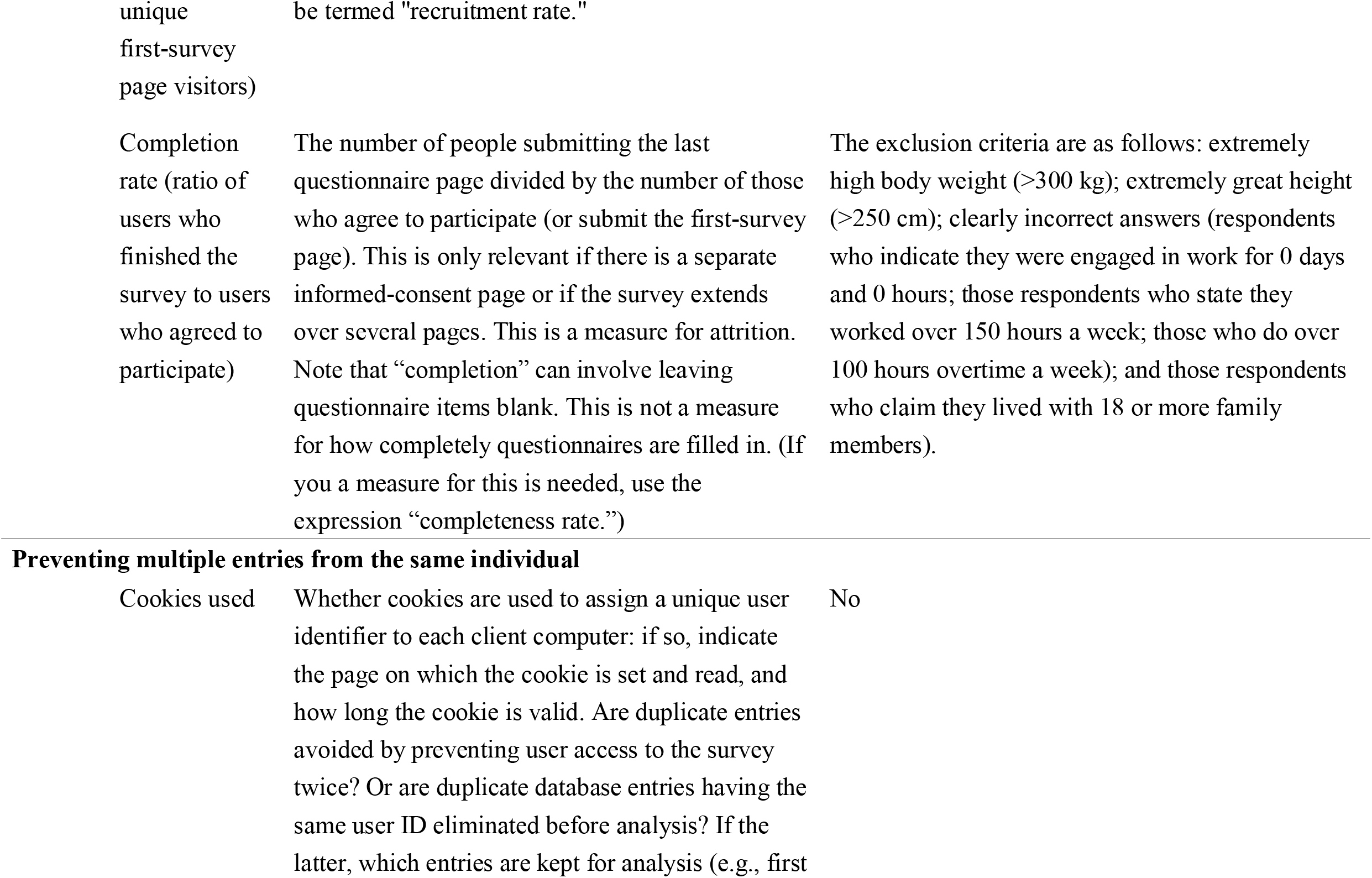
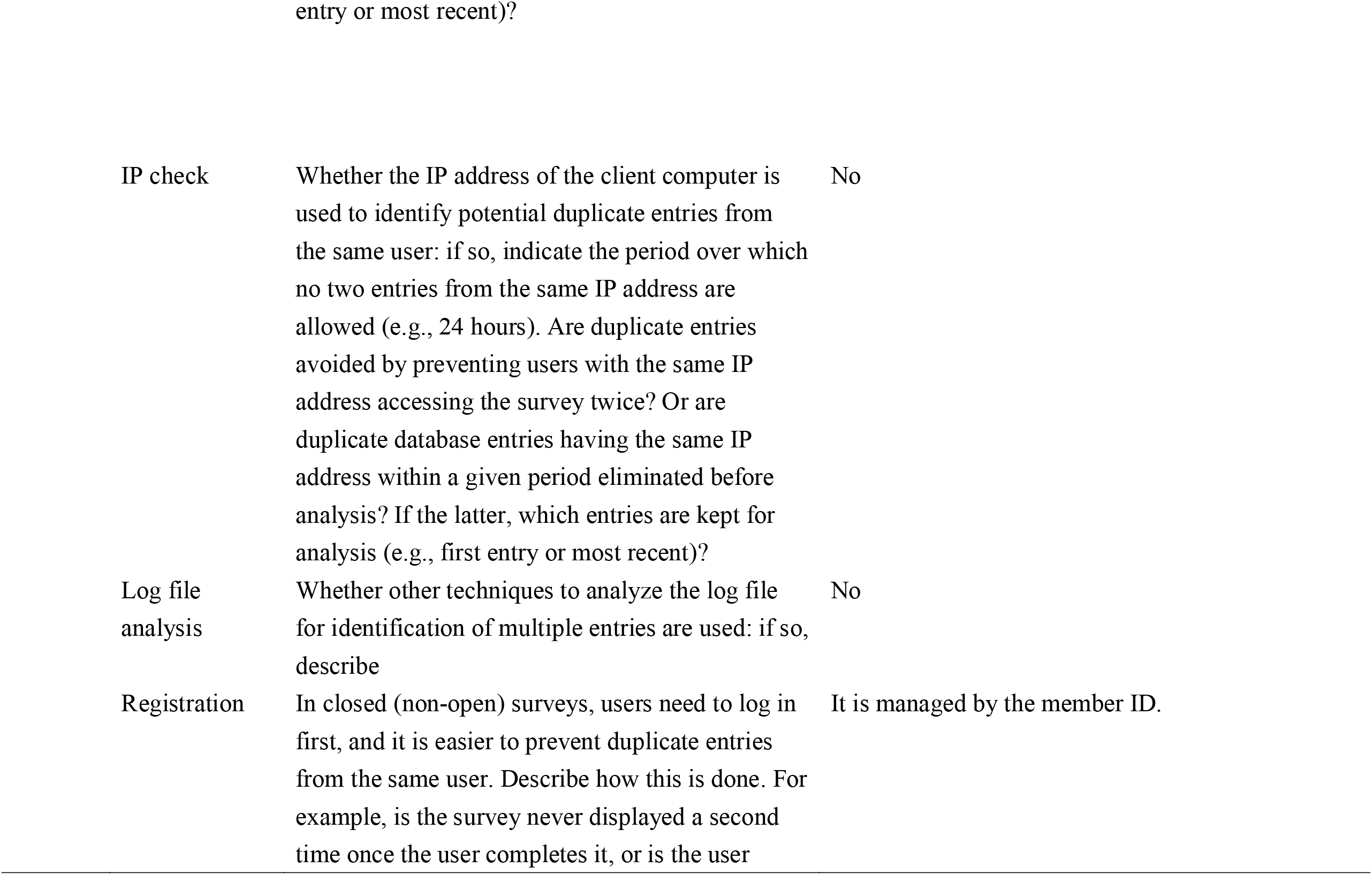
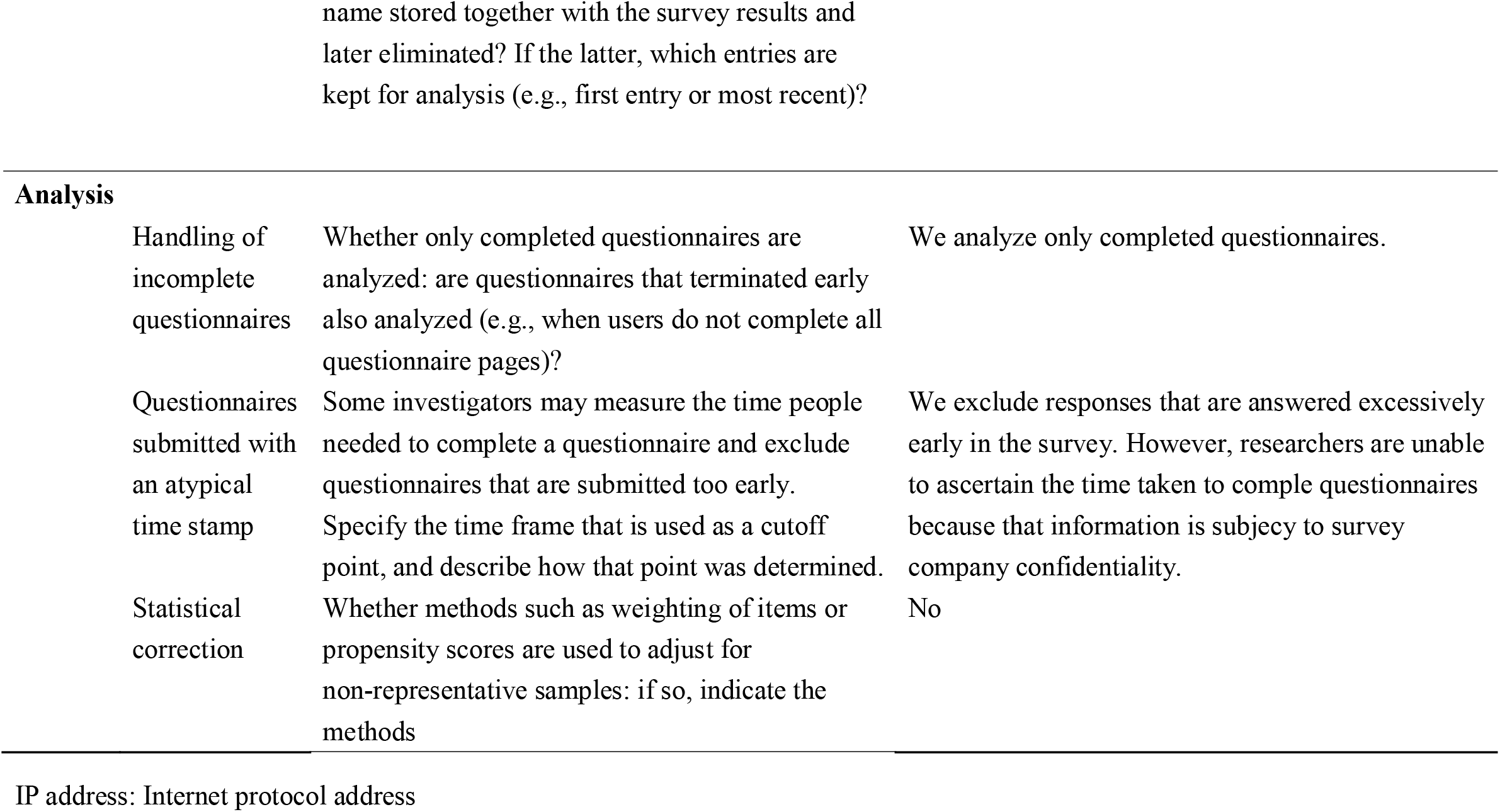
Checklist for Reporting Results of Internet E-Surveys (CHERRIES)

Table 3 presents specifics of the valid and invalid participants. The invalid responses comprised 1,244 (7.6%) men and 1,060 (7.8%) women. The proportion of invalid respondents ranged from 6.4% of women in their 30s to 11.2% of women aged over 65 years.

**Table 3.** Valid and invalid respondents stratified by sex and age category

| Age<br>category<br>(years) | Valid respondents |  | Invalid respondents |  |
| --- | --- | --- | --- | --- |
|  | Men<br>(15,201) | Women<br>(12,492) | Men<br>(1,244) | Women<br>(1,060) |
| 20–29 | 2,366 | 2,188 | 237 (9.1%) | 191 (8.0%) |
| 30–39 | 2,896 | 2,355 | 246 (7.8%) | 160 (6.4%) |
| 40–44 | 1,722 | 1,444 | 152 (8.1%) | 116 (7.4%) |
| 45–49 | 1,999 | 1,715 | 141 (6.6%) | 138 (7.4%) |
| 50–54 | 1,731 | 1,489 | 129 (6.9%) | 111 (6.9%) |
| 55–59 | 1,506 | 1,235 | 115 (7.1%) | 112 (8.3%) |
| 60–64 | 1,238 | 922 | 93 (7.0%) | 87 (8.6%) |
| ≥65 | 1,743 | 1,144 | 131 (7.0%) | 145 (11.2%) |

Table 4 shows the demographic characteristics and mental health–related indicators (K6 and work engagement score) for the valid and invalid participants. No differences were found between the groups for age, sex, and nationality. However, the invalid participants had statistically significantly higher psychological distress (*P* <0.001) and marginal significantly lower work engagement (*P* = 0.075) than the valid participants.

**Table 4.** Sociodemographic and mental health-related factors among valid and invalid respondents

|  | Valid respondents<br>(27,693) | Invalid respondents<br>(2,304) | *P* value |
| --- | --- | --- | --- |
| Age, mean (SD) | 45.8 (13.5) | 45.9 (14.3) | 0.530 |
| Sex (men), n (%) | 15,201 (54.9%) | 1,244 (54.0%) | 0.410 |
| Nationality (Japan), n (%) | 27,524 (99.4%) | 2,289 (99.3%) | 0.810 |
| K6, mean (SD) | 4.3 (4.8) | 4.7 (5.3) | <0.001 |
| WE, mean (SD) | 2.5 (1.4) | 2.4 (1.4) | 0.075 |

SD, standard deviation; K6, Kessler 6; WE, work engagement

Table 5 summarizes the characteristics of the valid participants by sex. The number and mean age of men were 15,201 (55%) and 46 years; those of women were 12,492 (45%) and 45 years. We observed that 0.6% of valid participants had differences between their biological and self-perceived sex. In all, 59% of men were married compared with 51% for women. Regarding employment status, 66% of men were permanent employees; 41% of women were permanent employees, and 34% of women had part-time employment. By industry, 21% of men and 10% of women worked in manufacturing; 8% of men and 21% of women worked in medical, health care and welfare; and 8% of men and 4% of women worked in the public sector. Regarding mental health status, 35% of men and 40% of women had a K6 score of 5 or higher, indicating mild psychological distress.

**Table 5.** Valid participant characteristics by sex

|  | Total<br>27,693 | Sex |  |
| --- | --- | --- | --- |
|  |  | Men<br>(15,201) | Women<br>(12,492) |
| Age, mean (SD) | 45.8 (13.5) | 46.3 (13.6) | 45.1 (13.4) |
| Gender, n (%) |  |  |  |
| Male | 15,159 (54.7%) | 15,120 (99.5%) | 39 (0.3%) |
| Female | 12,437 (44.9%) | 43 (0.3%) | 12,394 (99.2%) |
| Other | 97 (0.4%) | 38 (0.2%) | 59 (0.5%) |
| Nationality (Japanese), n (%) | 27,524 (99.4%) | 15,105 (99.4%) | 12,419 (99.4%) |
| Marital status, n (%) |  |  |  |
| Married | 15,241 (55.0%) | 8,928 (58.7%) | 6,313 (50.5%) |
| Unmarried | 8,362 (30.2%) | 4,497 (29.6%) | 3,865 (30.9%) |
| Divorced or bereaved | 4,090 (14.8%) | 1,776 (11.7%) | 2,314 (18.5%) |
| Household income (yen), n (%) |  |  |  |
| <4 million | 6,926 (25.0%) | 3,100 (20.4%) | 3,826 (30.6%) |
| 4 million–6 million | 6,919 (25.0%) | 3,918 (25.8%) | 3,001 (24.0%) |
| 6 million–8 million | 5,478 (19.8%) | 3,138 (20.6%) | 2,340 (18.7%) |
| 8 million–10 million | 3,814 (13.8%) | 2,232 (14.7%) | 1,582 (12.7%) |
| 10 million–12 million | 1,871 (6.8%) | 1,163 (7.7%) | 708 (5.7%) |
| >12 million | 2,685 (9.7%) | 1,650 (10.9%) | 1,035 (8.3%) |
| Employment status, n (%) |  |  |  |
| Self-employed | 2,546 (9.4%) | 1,654 (10.9%) | 942 (7.5%) |
| Company executive | 1,595 (5.8%) | 1,213 (8.0%) | 382 (3.1%) |
| Full-time employee | 15,240 (55.0%) | 10,064 (66.2%) | 5,176 (41.4%) |
| Part-time work | 5,217 (18.8%) | 936 (6.2%) | 4,281 (34.3%) |
| Dispatched employee | 574 (2.1%) | 158 (1.0%) | 416 (3.3%) |
| Contract employees | 1,747 (6.3%) | 888 (5.8%) | 859 (6.9%) |
| Freelance | 207 (0.7%) | 33 (0.2%) | 174 (1.4%) |
| Other | 517 (1.9%) | 255 (1.7%) | 262 (2.1%) |
| Industry category, n (%) |  |  |  |
| Agriculture and forestry | 261 (0.9%) | 172 (1.1%) | 89 (0.7%) |
| Fisheries | 22 (0.1%) | 13 (0.1%) | 9 (0.1%) |
| Mining and quarrying of stone<br>and gravel | 29 (0.1%) | 23 (0.2%) | 6 (<0.1%) |
| Construction | 1,399 (5.1%) | 980 (6.4%) | 419 (3.4%) |
| Manufacturing | 4,432 (16.0%) | 3,246 (21.4%) | 1,186 (9.5%) |
| Electricity, gas, heat supply<br>and water | 391 (1.4%) | 295 (1.9%) | 96 (0.8%) |
| Information and<br>communications | 1,369 (4.9%) | 982 (6.5%) | 387 (3.1%) |
| Transport and postal services | 1,221 (4.4%) | 909 (6.0%) | 312 (2.5%) |
| Wholesale and retail trade | 2,887 (10.4%) | 1,292 (8.5%) | 1,595 (12.8%) |
| Finance and insurance | 1,197 (4.3%) | 549 (3.6%) | 648 (5.2%) |
| Real estate and goods rental<br>and leasing | 702 (2.5%) | 452 (3.0%) | 250 (2.0%) |
| Scientific research,<br>professional and technical<br>services | 788 (2.8%) | 494 (3.2%) | 294 (2.4%) |
| Accommodations, eating and<br>drinking services | 968 (3.5%) | 326 (2.1%) | 642 (5.1%) |
| Living-related and personal<br>services and amusement<br>services | 749 (2.7%) | 294 (1.9%) | 455 (3.6%) |
| Education and learning support | 1,718 (6.2%) | 719 (4.7%) | 999 (8.0%) |
| Medical, health care and<br>welfare | 3,775 (13.6%) | 1,218 (8.0%) | 2,557 (20.5%) |
| Compound services | 267 (1.0%) | 154 (1.0%) | 113 (0.9%) |
| Services | 2,835 (10.2%) | 1,469 (9.7%) | 1,366 (10.9%) |
| Public sector | 1,645 (5.9%) | 1,165 (7.7%) | 480 (3.8%) |
| Unlabeled | 1,038 (3.7%) | 449 (3.0%) | 589 (4.7%) |
| K6, mean (SD) | 4.3 (4.8) | 4.0 (4.7) | 4.6 (4.9) |
| K6 (5 and more), n (%) | 10,328 (37.3%) | 5,285 (34.8%) | 5,043 (40.4%) |
| K6 (10 and more), n (%) | 3,967 (14.3%) | 2,023 (13.3%) | 1,944 (15.6%) |
| K6 (13 and more), n (%) | 1,916 (6.9%) | 952 (6.3%) | 964 (7.7%) |
| WE, mean (SD) | 2.5 (1.4) | 2.4 (1.4) | 2.5 (1.4) |

444 K6, Kessler 6; WE, work engagement; SD, standard deviation

## Discussion

We conducted an Internet-based survey of workers in Japan in March 2022. We reported the details of the study based on the CHERRIES. Our sample had the same sex, age, and regional composition as the Japanese working population.

With the development of information and communication technology, questionnaire surveys are increasingly being conducted through the Internet (5). Web-based surveys have been used more frequently following COVID-19. In an emergency situation, such as a pandemic, investigations need to be quickly performed. Thus, Internet surveys are being increasingly carried out using panels registered with research firms.

In this study, we surveyed panel registrants. We took steps to exclude participants who made dishonest responses. Among the invalid respondents, we found no bias regarding sex, age, or nationality. We conducted our survey only in Japanese language and foreign nationals proficient in that language. Invalid participants exhibited higher levels of psychological distress and lower work engagement than the valid ones. Individuals with poor mental health are subject to greater fatigue and have lower attention spans, which increase the likelihood of invalid responses (18). Alternatively, giving inaccurate answers may be associated with psychological distress and poor work engagement (19).

Among our valid participants, 0.6% indicated differences with respect to sex and gender. In one Japanese survey conducted in 2019 (20), 0.7% of respondents answered no to the question “Do you see your gender now as the same as your sex at birth?” That survey was conducted on a general population (of whom 80% were workers); however, the proportion of individuals with differences regarding sex and gender was almost the same as in our survey. We found that non-Japanese participants accounted for 0.6% of the total. According to a nationwide survey, the number of foreign workers in Japan was 1,727,221 as of October 31, 2021, and they accounted for 2.5% of the overall workforce (21). The low percentage of non-Japanese participants in our survey suggests that few of them were registered with that Internet panel.

We found that the proportion of married participants was 58.7% for men and 50.5% for women. According to the national census of 2020 (22), 57.4% of married people in Japan were men and 54.0% were women; in 2015, the figures were 58.9% and 55.2%, respectively; thus, a decrease occurred for both sexes. We found that among our valid participants, the proportion of married people was higher for men and lower for women than in the general population (7). When asked if obstacles existed to getting married within the next year, one study found that over 40% of both men and women cited wedding costs and funding necessary to set up a new life (23). Financial reasons may deter unemployed men from getting married. By contrast, women may defer marriage through having to stop work following marriage or childbirth. According to one study, the average household income in Japan in 2020 was 5,643,000 yen (24). We found that 53.8% of men and 45.4% of women had annual household incomes of 6 million yen or more; thus, our participants had similar income levels to the overall worker population in Japan.

Regarding employment status, our participants displayed similar proportions to those identified in a nationwide statistical survey in Japan (24), indicating that our sampling was appropriate. In the national survey, 11.4% of men and 8.6% of women were self-employed; in our study, those figures were 10.9% and 7.5%, respectively.

Among workers excluding self-employed individuals and company executives, the proportion of full-time employees was 77.8% for men and 46.6% for women in that national survey; in our study, those figures were 81.6% and 46.3%, respectively. Similar findings emerged with respect to industry. Manufacturing accounts for the largest number of employees: the national survey found that 20.5% of men and 10.6% of women worked in that sector; our results indicated 21.4% of men and 9.5% of women, respectively. Our study was conducted approximately 2 years after COVID-19 began. We found that medical, health care and welfare accounted for 8.0% of men and 20.5% of women; those figures do not markedly differ from the 6.4% and 23.1%, respectively, in the national survey. Thus, we believe that COVID-19 did not prevent health workers from participating in our study.

We used the K6 score for depression and anxiety to assess mental health status. A K6 score of 10 or higher indicates experiencing psychological distress equivalent to mood or anxiety disorders (25). The proportion of our participants with a score of 10 or higher (13.3% for men and 15.6% for women) was exactly the same as observed in the national survey (25). Conversely, the proportions of participants scoring 5 or higher (moderate risk of depression and anxiety) were 29.6% for men and 34.8% for women in the national survey in 2019; we recorded 34.8% and 40.4%, respectively. That national survey was based on data from before COVID-19. Two years after the pandemic began, the levels of depressive and anxiety disorders were not so high; however, many people still apparently experienced mild anxiety. Work engagement is a positive aspect of mental health; it was found to be 2.4 for men and 2.4 for women in a large Japanese survey conducted after the start of the pandemic (26); in this study we recorded figures of 2.4 and 2.5, respectively.

In conclusion, the present study describes in detail the protocol of a baseline survey for a cohort study of the Japanese workforce population. We stratified sex, age, and region such that our population ratios matched the nationwide situation. We also reported the details of the study based on the CHERRIES. Our results indicate that the sampling was adequate in terms of employment status and industry. We plan to use this database to develop benchmarks related to occupational health and safety.

## Data Availability

All data produced in the present study are available upon reasonable request to the authors.

## Acknowledgments

The current members of the W2S-Ohpm Study, in alphabetical order, are as follows: Akiko Matsuyama, Asumi Yama, Ayaka Yamamoto, Ayana Ogasawara, Hideki Fujiwara, Juri Matsuoka, Kakeru Tsutsumi, Kazufumi Matsuyama, Kenta Moriya, Kiminori Odagami, Koji Mori, Kosuke Sakai, Masako Nagata, Miho Omori, Mika Kawasumi, Mizuho Inagaki, Naoto Ito, Rina Minohara, Shunusuke Inoue, Suo Taira, Takahiro Mori, Tomohisa Nagata (present chairperson of the study group), and Tomoko Sawajima. All members are affiliated with the University of Occupational and Environmental Health, Japan. We thank Edanz (https://www.edanz.com/ac) for editing a draft of this manuscript.

## Authors contribution

TN, KO and KM, study design; TN, KO, NA, MN and KM, data collection; ST and TN, data analysis; ST, draft of manuscript; All authors have reviewed, edited and approved the final manuscript.

